# EPICOVID19: Psychometric assessment and validation of a short diagnostic scale for a rapid Covid-19 screening based on reported symptoms

**DOI:** 10.1101/2020.07.22.20159590

**Authors:** Luca Bastiani, Loredana Fortunato, Stefania Pieroni, Fabrizio Bianchi, Fulvio Adorni, Federica Prinelli, Andrea Giacomelli, Gabriele Pagani, Stefania Maggi, Caterina Trevisan, Marianna Noale, Nithiya Jesuthasan, Aleksandra Sojic, Carla Pettenati, Massimo Andreoni, Raffaele Antonelli Incalzi, Massimo Galli, Sabrina Molinaro

## Abstract

**Background:** Confirmed COVID-19 cases have been registered in more than two hundred countries and regions and of July 28 over 16 million cases of COVID-19, including 650805 deaths, have been reported to WHO. The number of cases changes quickly and varies depending upon which source you use to track, so in the current epidemiological context, the early recognition is critical for the rapid identification of suspected cases (with SARS-CoV-2 infection-like symptoms and signs) to be immediately subjected to quarantine measures. Although surveys are widely used for identifying COVID-19 cases, outcomes and associated risks, no validated epidemiological tool exists for surveying SARS-CoV-2 infection in the population so far.

**Methods:** Our study is the phase II of the EPICOVID19 Italian national survey, launched in April 2020 including a national convenience sample of 201121 adults, who voluntarily filled the EPICOVID19 questionnaire. The phase II questionnaire was mailed to all subjects who underwent tests for COVID-19 by nasopharyngeal swab (NPS) and who accepted to be involved in the second phase of the study, focused on the results reported for NPS and/or serological IgG/IgM tests. We evaluated the capability of the self-reported symptoms collected through the EPICOVID19 questionnaire to discriminate the COVID-19 among symptomatic subjects, in order to identify possible cases to undergo instrumental measurements and clinical examinations. We defined a method for the identification of a total score and validated it with reference to the serological and molecular clinical diagnosis, using four standard steps: identification of critical factors, confirmation of presence of latent variable, development of optimal scoring algorithm and validation of the scoring algorithm.

**Findings:** 2703 subjects [66% response rate] completed the Phase II questionnaire. Of 2703 individuals, 694 (25.7%) were NPS(+) and of these 84 (12.1% of the 694 NPS(+)) were asymptomatic. In the individuals who performed serological testing, of the 472 who did IgG(+) and 421 who did IgM(+), 22.9% and 11.6% tested positive, respectively. Among IgG(+) 1 of 108 subjects was asymptomatic (0.9%) while 5/49 subjects among IgM(+) were asymptomatic (10.2%). Compared with NPS(-), among NPS(+) subjects there was a higher rate for Fever (421 [60.7%] vs 391[19.5%]; p<0.0001), Loss of Taste and/or Smell (365 [52.6%] vs 239 [11.9%]; p<0.0001) and Cough (352 [50.7%] vs 580 [28.9%]; p<0.0001). Also for other symptoms the frequencies were significantly higher in NPS(+) subjects than in NPS(-) ones (p<0.001). Among groups with serological tests, the symptoms with higher percentages in the subjects IgG(+) were Fever (65 [60.2%] vs 43[11.8%]; p<0.0001) and Pain in muscles, bones, joints (73 [67.6%] vs 71 [19.5%]; p<0.0001). For the COVID-19 self-reported symptoms items, exploratory (proportion variance explained [89.9%]) and confirmatory factor analysis results (SMSR 0.072; RMSEA 0.052) highlights the presence of one latent variable (factor) underlying the symptoms. We define the one-factor solution as EPICOVID19 diagnostic scale and optimal score for each items was identified: Respiratory problems (1.03), Chest pain (1.07), Loss of Taste and/or Smell (0.97) and Tachycardia (palpitations) (1.05) were the most important symptoms.

The cut-off score was 2.56 (Sensitivity 76.56%; Specificity 68.24%) in NPS(+) and 2.59 (Se 80.37; Sp 80.17) in IgG(+) subjects.

**Interpretation:** We developed a short diagnostic scale to detect subjects with symptoms potentially associated with COVID-19 among a wide population. Early recognition screening and rapid diagnosis are essential to prevent transmission and provide supportive care in a timely manner and our score supports the potential for identifying individuals who need to seek immediate clinical evaluation. Although these results are referred to the Italian pandemic period, this short diagnostic scale could be optimised and tested as a screening tool in other similar pandemic contexts.

## Introduction

SARS Corona Virus 2 has led to a global pandemic (on July 28 reports over 16 million cases and 650805 deaths across more than 200 countries, WHO, Johns Hopkins Center for Health Security) ^1,2^. Besides this immediate human toll there are readily acknowledged and potentially long-lasting effects on global economies, politics, health and privacy policies at many levels that will extend beyond the development of vaccines and treatments. The rapid spread of the disease, COVID-19, and its seemingly high degree of variability in its presentation among individuals has led to a level of clinical and scientific focus not previously seen and encompassing both traditionally reviewed and pre-print publications and resources. Collaborative groups are being formed at the local, regional, national and international levels to address both patient data collection/aggregation and analysis in ways that may change the way research is carried out in the future^3^. To enable these efforts to be both effective and productive is the need for the data to be evaluated as to its suitability for inclusion in these activities while still recognizing that what we understand about COVID-19 is much less than what we do not understand^4^.

Because of the far-reaching scope of the pandemic, we are already confronting:

1. Need to implement individual testing at a level far above current capacities to optimize individual treatment, assess disease spread, anticipate potential strains on healthcare resources and personnel^5^.
2. Need for improvements in available testing, both nasopharyngeal swabs (NPS) and antibody detection, i.e. accuracy, specificity and sensitivity to enable reliable evaluation and interpretation of data for use in clinical care and policy decisions^6^.
3. Need to harmonize clinical observations and definitions to support development of guidelines, prognostic and diagnostic indicators and to develop a comprehensive understanding of the disease and critical factors that differentiate patient susceptibility, presentation of the disease and response to treatment^7,8^.

The use of online surveys can greatly enhance access to broader populations in a cost-effective manner and optimize both screening for individuals who may need immediate care as well as provide an approach to 3) above. A cross-sectional national survey, EPICOVID19, was launched on April 13, 2020 and received more than 200000 responses^9^. The survey, which represents the Phase I of the study, was promoted using social media (Facebook, Twitter, Instagram, Whatsapp), press releases, internet pages, local radio and TV stations, and institutional websites that called upon volunteers to contact the study website. The inclusion criteria were: age of >18 years; access to a mobile phone, computer, or tablet with internet connectivity; and on-line consent to participate in the study.

The aim of our study is to assess the capability of the self-reported symptoms collected through the EPICOVID19 questionnaire to discriminate the COVID-19 among symptomatic subjects, in order to identify possible COVID-19 cases to undergo instrumental measurements and clinical examinations (Phase II of the study). The final objectives are to propose a method for the development of a total score for the self-reported symptoms in the EPICOVID19 questionnaire and to validate the scoring method with reference to the molecular and serological clinical diagnosis.

## Methods

### Study design and participants

This current study is the Phase II of the EPICOVID19 national survey^9^. The Figure 1 shows the overview of EPICOVID19 two-phase study. The Phase I questionnaire investigated six areas through 38 questions (1: Socio-demographic characteristics; 2: Clinical evaluation; 3: Personal characteristics and health status; 4: Housing conditions; 5: Lifestyle; 6: Behaviours after the lockdown).

**Figure 1.**
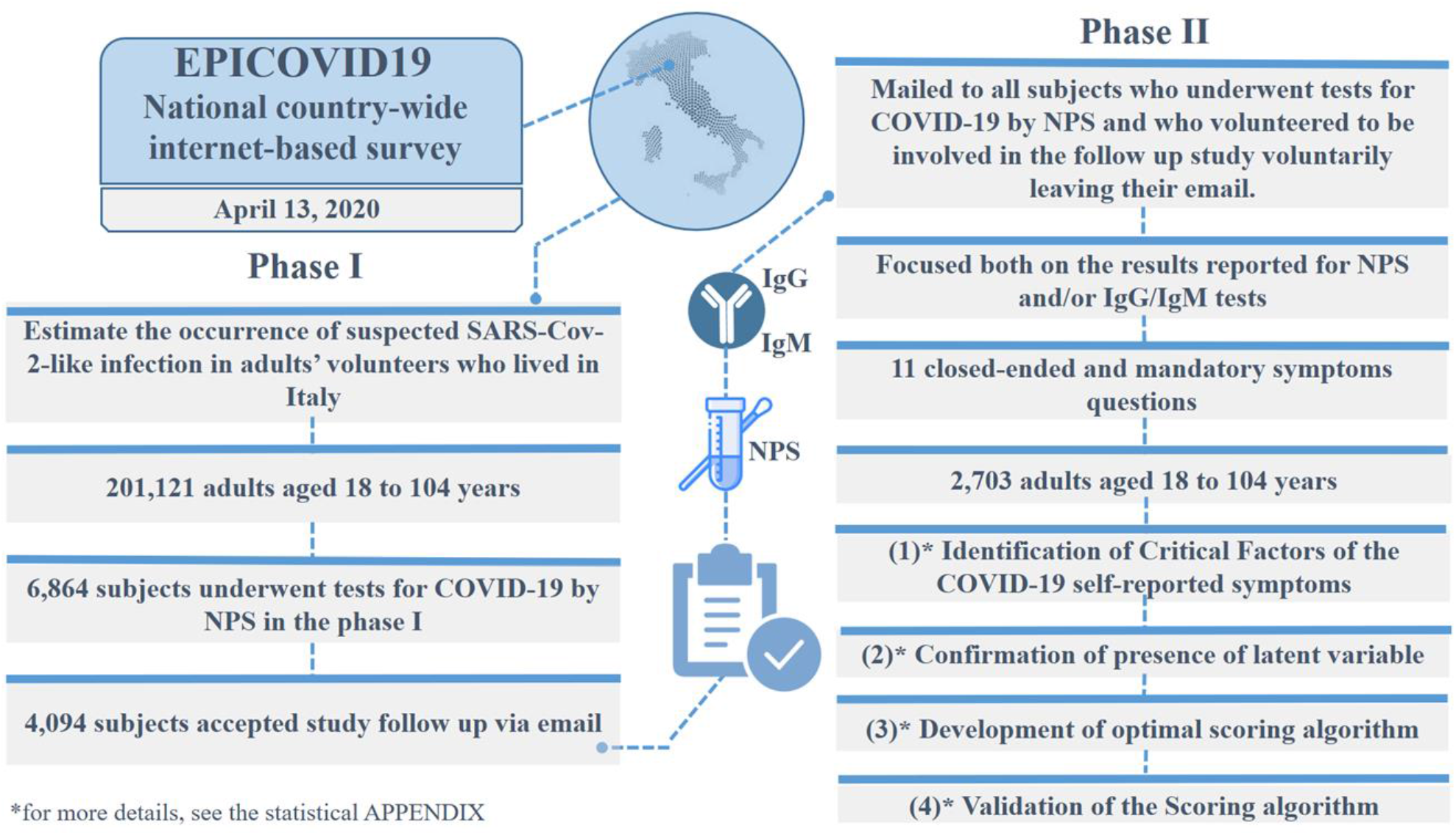
Overview of EPICOVID19 two-phase study.

The Phase II questionnaire was mailed to all subjects who underwent testing for COVID-19 by NPS, and who volunteered to be involved in the follow up study in their Phase I response. Phase II focused on results reported for NPS and/or serological IgG/IgM tests and self-reported symptoms with the aim to better identify both symptomatic and asymptomatic cases of SARS-CoV-2 infections^10^.

Phase II was implemented using an open source statistical survey framework PHP based, LimeSurvey (vers. 3.17), distributed under the GNU General Public License. In Phase II, responses to 11 questions were required that covered administration of NPS and/or serological tests and on the time elapsed between observed/reported symptoms and clinical examination (NPS and/or antibodies IgG and IgM) (appendix p 1-8). Of the 6864 subjects who underwent testing for COVID-19 by NPS in the phase I, 4094 subjects were invited by e-mail to complete the Phase II questionnaires online. Of these, 38 email invitations were not delivered, e.g. wrong address, mailbox full, host or domain name not found, etc.; 101 individuals did not participate by refusing to provide consent; and 1252 individuals who received the email did not proceed to complete the questionnaire. A total of 2703 subjects (66% response rate) completed the phase II survey. We compared the characteristics of respondents (2703) and non-respondents (4161). Respondents and non-respondents to the Phase II survey appeared similar with respect to gender, age and the perception of their own health as well as their self-reported comorbidities; the details comparing these two groups of subjects are included in appendix p 9. The resulting data of 2703 subjects who completed the Phase II were linked to the results of the self-reported symptoms of Phase I EPICOVID19 questionnaire, which included questions on the presence of 11 symptoms.

### Statistical analysis

We analysed the self-reported symptoms collected in the survey to define a method for the construction of a total score and to validate the scoring method with reference to the serological and molecular clinical diagnosis, using four standard questionnaire validation steps:

1. **Identification of Critical Factors:** We determined the factorial structure of the COVID-19 self-reported symptoms items using exploratory factor analysis (EFA) followed by confirmatory factor analysis (CFA). EFA and parallel analysis (PA) were performed to identify the performance of specific symptoms (loadings) and to define the number of factors underlying these loadings.
2. **Confirmation of presence of latent variable**: We carried out CFA via Structural Equation Modelling (SEM) to confirm the presence of one latent variable (factor) underlying the 11 symptoms chosen to identify COVID-19. Several goodness-of-fit criteria were used: (i) Standardized Root Mean Square Residual (SRMR) and (ii) Root Mean Square Error of Approximation (RMSEA) (not higher than 0.10); (iii) Comparative fit index (CFI) and (iv) Tucker-Lewis index (TLI) (not less than 0.90).
3. **Development of optimal scoring algorithm**: We developed an optimal scoring algorithm using Homogeneity Analysis (Multiple Correspondence Analysis). Through the HOMALS procedure we replaced specific dichotomous responses, i.e. Yes/No, with categorical quantifications: the resulting score is the sum of the subject’s symptom responses, once they are re-coded by category quantifications.
4. **Validation of the Scoring algorithm**: We validated the score using an external objective criterion based on receiver operating characteristics (ROC) analysis, in order to evaluate the COVID-19 symptoms score performance in distinguishing symptomatic individuals. With the aim of discriminating COVID-19 cases, sensitivity, specificity, and Youden’s index were computed with two reference standards: a) subjects with positive NPS vs subjects with negative NPS; b) subjects with serological IgG(+) vs subjects with IgG(-). The overall predictive performance was evaluated by the AUC (Area Under the Curve).

All statistical analyses were carried out using R software (version 3.6.3). The details of the performed statistical analysis are reported in appendix p 10-12.

## Results

The characteristics of the 2703 subjects, as supplied by those who completed the phase II survey, their NPS, IgG and IgM results are shown in table 1. The sample was predominantly women (68.1%) with the average age being 49 ± 15.0 and 52 ± 14.1 years for women and male individuals, respectively.

**Table 1.** Self-reported characteristics from the survey analysed using tests’ results for SARS-CoV-2 infection.

|  | Tested for SARS-CoV-2 |  |  |  |  |  |  |  |  |
| --- | --- | --- | --- | --- | --- | --- | --- | --- | --- |
|  | NASOPHARYNGEAL SWAB |  |  | ANTIBODIES IgG |  |  | ANTIBODIES IgM |  |  |
|  | (2703) |  |  | (472) |  |  | (421) |  |  |
|  | Tested positive | Tested negative | p value | Tested positive | Tested negative | p value | Tested positive | Tested negative | p value |
| Number | 694 (25.7%) | 2009 (74.3%) |  | 108 (22.%) | 364 (77.1%) |  | 49 (11.6%) | 372 (88.4%) |  |
| Women | 440 (63.4%) | 1401 (69.7%) | 0.001 | 61 (56.5%) | 258 (70.9%) | 0.005 | 25 (51.0%) | 260 (69.9%) | 0.008 |
| (%) |  |  |  |  |  |  |  |  |  |
| Age (years) media e dev standard | 55.5 ±18.06 | 47.55 ±12.81 | <0.0001 | 48.8 ±11.74 | 45.5 ±11.49 | 0.009 | 50.6 ±10.56 | 45.8 ±11.69 | 0.008 |
| Answered questions on symptoms (n) |  |  |  |  |  |  |  |  |  |
| (S1) Fever, with temperatures above 37.5 ° C for at least three consecutive days | 421 (60.7) | 391 (19.5%) | <0.0001 | 65 (60.2%) | 43 (11.8%) | <0.0001 | 28 (57.1%) | 68 (18.3%) | <0.0001 |
| (S2) Cough | 352 (50.7%) | 580 (28.9%) | <0.0001 | 63 (58.3%) | 76 (20.9%) | <0.0001 | 26 (53.1%) | 95 (25.5%) | <0.0001 |
| (S3) Sore throat and/or cold | 232 (33.4%) | 756 (37.6%) | 0.048 | 46 (42.6%) | 132 (36.3%) | 0.233 | 16 (32.7%) | 135 (36.3%) | 0.617 |
| (S4) Headache | 313 (45.1%) | 703 (35.0%) | <0.0001 | 61 (56.5%) | 96 (26.4%) | <0.0001 | 23 (46.9%) | 117 (31.5%) | 0.031 |
| (S5) Pain in muscles, bones, joints | 360 (51.9%) | 572 (28.5%) | <0.0001 | 73 (67.6%) | 71 (19.5%) | <0.0001 | 27 (55.1%) | 98 (26.3%) | <0.0001 |
| (S6) Loss of taste and / or smell | 365 (52.6%) | 239 (11.9%) | <0.0001 | 66 (61.1%) | 29 (8.0%) | <0.0001 | 21 (42.9%) | 55 (14.8%) | <0.0001 |
| (S7) Respiratory difficulty (sense of breathlessness at rest), | 179 (25.8%) | 249 (12.4%) | <0.0001 | 21 (19.4%) | 28 (7.7%) | <0.0001 | 7 (14.3%) | 37 (9.9%) | 0.350 |
| (S8) Chest pain (sternum pain) | 136 (19.6%) | 251 (12.5%) | <0.0001 | 26 (24.1%) | 25 (6.9%) | <0.0001 | 7 (14.3%) | 37 (9.9%) | 0.350 |
| (S9) Tachycardia (Palpitations) | 120 (17.3%) | 237 (11.8%) | <0.0001 | 24 (22.2%) | 27 (7.4%) | <0.0001 | 10 (20.4%) | 31 (8.3%) | 0.007 |
| (S10) Gastrointestinal complaints (diarrhoea, nausea, vomiting) | 289 (41.6%) | 452 (22.5%) | <0.0001 | 54 (50.0%) | 65 (17.9%) | <0.0001 | 17 (34.7%) | 87 (23.4%) | 0.084 |
| (S11) Conjunctivitis (red eyes) | 111 (16.0%) | 221 (11.0%) | 0.001 | 24 (22.2%) | 35 (9.6%) | 0.001 | 11 (22.4%) | 40 (10.8%) | 0.018 |

Notes. Numbers are mean ± SD for continuous variables (independent t-test) and frequency (%) for the categorical ones (Chi square test)

In the total sample of 2703 individuals, 694 (25.7%) were NPS(+) and of these 84 (12.1% of the 694 NPS(+)’s) were asymptomatic.

Testing statistics: for the subgroup of individuals who performed serological testing, of the 472 who did IgG(+) and 421 who did IgM(+), 22.9% and 11.6% tested positive, respectively. Among IgG(+) 1 of 108 subjects was asymptomatic (0.9%) while 5/49 subjects among IgM(+) were asymptomatic (10.2%). For subjects with NPS(+), the average number of days between initial symptoms and the day of the swab execution was 9.3 ± 9.4 (median 7 days, IQR 3-7). For subjects IgG(+) the average number of days between initial symptoms and the day of the serological test execution was 36.1 ± 15.1 (median 36.5 days, IQR 28-47). For subjects IgM(+) the average number of days from initial symptoms to the day of the serological test execution was 26.1 ± 17.9 (median 28.0 days, IQR 4-40). The frequency of the eleven symptoms reported by the three groups (NPS, IgG, IgM tested) was similar among men and women. In NPS(+) group, women reported higher percentages than male for the Sore throat and/or cold and/or Tachycardia (Palpitations) symptoms only. In the IgG(+), males reported higher frequencies for headaches only, while in the IgM(+) group the females showed the lower frequency of symptoms related to Conjunctivitis.

The frequency of symptoms among NPS(+) subjects (table 1) ranged from a low rate of observation, e.g. Tachycardia (palpitations) (S9: 17.3%) and Conjunctivitis (S11: 16.0%), to a high observation rate, e.g. Fever (S4: 60.7%) and OT-D (S6: 52.6%). For all symptoms, apart from Headache, the frequencies were significantly higher in NPS(+) subjects than in NPS(-) ones (p<0.001). For the subgroup of individuals who also performed serological tests, the symptoms with higher percentages among the subjects tested positive were Fever (IgG(+) 60.2%; IgM(+) 57.1%) and Pain in muscles, bones, joints IgG(+) 67.6%; IgM(+) 55.1%). In the IgG serological group, no difference was observed in the percentages of Sore throat and/or cold (S3) symptoms, while for Respiratory difficulty (S7), Chest pain (S8) and Gastrointestinal symptoms (S10), the percentages in the IgM group were the same.

The EFA performed by Principal-Component Factors and Horn’s PA methods statistics pointed out one factor. Eigenvalues, descriptive indices, and goodness-of-fit indices of cumulative percentages of explained data variability obtained through EFA are displayed in table 2. Principal-Component Factors highlight only one factor with 89.9% of proportion of explained variability, while Horn’s PA method identifies two factors with eigenvalues greater than 1.0, with 49.8% and 10.3% of proportion of explained variability, respectively.

**Table 2.** **Descriptive and Goodness of fit dimensionality indices from EFA of the eleven EPICOVID19 symptoms items in 2703 subjects, using Principal-Component Factors and Horn’s Parallel Analysis methods with eigenvalue 1**.

| Exploratory factor analysis |  |  |  |  |  |  |
| --- | --- | --- | --- | --- | --- | --- |
| Principal-Component Factors |  |  |  | Horn's Parallel Analysis |  |  |
| Factor | Eigenvalue | Proportion of explained variability | Cumulative explained variability | Eigenvalue | Proportion of explained variability | Cumulative explained variability |
| 1 | 5.00 | 89.9% | 89.9% | 5.48 | 49.8 | 49.8% |
| 2 | - | - | - | 1.14 | 10.3 | 60.1% |

Based on a priori determined cut-off value the factor-loading greater than 0.35 was maintained. The factors loading rule of one factor solution extracted by Principal-Component Factors is available in appendix p 13. Then, the dimensionality indices of the one-factor solution, as the high variance explained by the factor (89.9%), confirms the presence of one latent variable underlying COVID-19 symptoms items. Therefore, we define the one-factor solution as “EPICOVID19 diagnostic scale” (EPICOVID19 DS). On the basis of the CFA, results confirmed the latent construct as uni-dimensional and how the variables contributed to EPICOVID19 DS. The figure 2 shows the values of the standardized factor loadings for the one factor model. The magnitude of each factor loading higher than 0.4 indicates the importance of the corresponding item to EPICOVID19 DS. For example, Pain in muscles, bones, joints were the most important variable with a value equal to 0.814. The other variables with optimal specific validity index were Respiratory difficulty (sense of breathlessness at rest (0.688), Loss of taste and/or smell (0.724) and Gastrointestinal complaints with item-factor correlations equal 0.737. The lowest values were observed for the Sore throat and/or cold and Conjunctivitis items, 0.537 and 0.557, respectively. The goodness of fit (SMSR, RMSEA) of the EPICOVID19 DS was acceptable because two indexes were lower than 0.10 (SMSR 0.072; RMSEA 0.052; CFI 0.977; TLI 0.971). Finally, we compute CFA indexes to measure internal validity of the model (appendix p 14).

**Figure 2.**
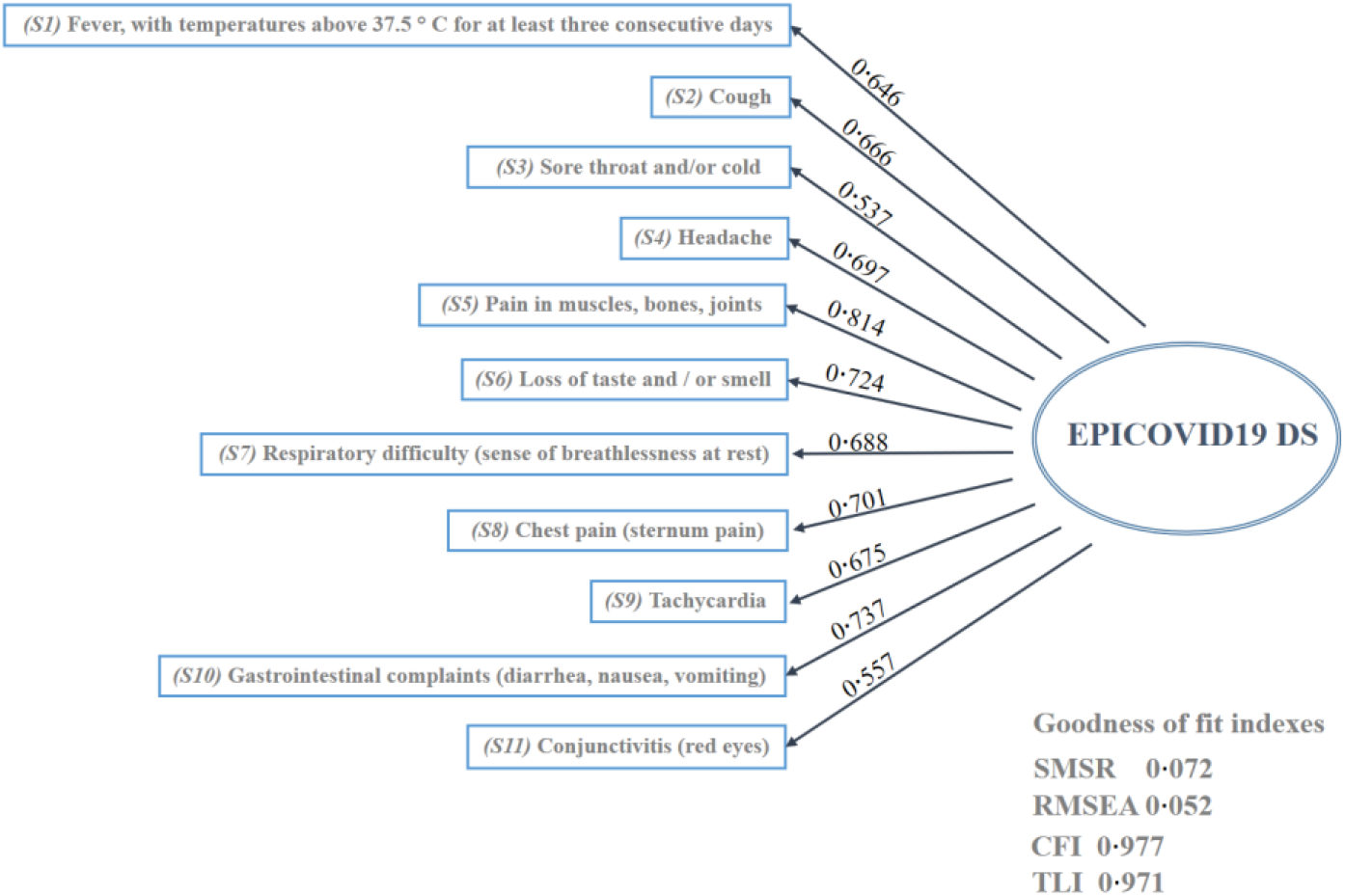
**Standardized factor loadings and goodness of fit indexes for one factor model. The goodness of fit indexes (SMSR, RMSEA) of the** “**EPICOVID19 DS”**

Given the successful uni-dimensionality testing of EPICOVID19 DS, optimal scaling via Homogeneity Analysis was undertaken. The optimal score proposed was extracted from the HOMALS procedure (single-factor measurement) and for each subject, the computed optimal score was obtained summing the category quantifications of the screening questionnaire item responses. Cronbach (0.88) and Greenacre (78%) indices confirm the uni-dimensionality found through EFA and CFA. The HOMALS optimal category quantifications of the EPICOVID19 symptoms variable are summarized in table 3. Columns show the binary options (YES-NO), while rows show the different symptoms. The HOMALS category quantifications were scaled so that the score obtained from the sum of responses, varies from 0 (if a subject answered NO to all the symptoms) to 10 (if a subject answered YES to all the symptoms). These values are shown in the last column of table 3. An example of the resulting score calculation is reported as follows. If the subject response pattern with respect to symptoms is: YES, NO, YES, NO, NO, YES, YES, NO, NO, NO, YES, the re-coded response pattern is: 0.80, 0, 0.64, 0, 0, 0.97, 1.03, 0, 0, 0, 0.88 and subject optimal score is: 0.8 + 0+ 0.64+0+ 0+ 0.97+ 1.03+ 0+ 0+ 0+ 0.88 = 4.2.

**Table 3.** Multiple Correspondence Analysis optimal weights recording of the EPICOVID-19 DS.

|  | HOMALS category quantifications |  | Recoded HOMALS category quantifications |  |
| --- | --- | --- | --- | --- |
|  | No | Yes | No | Yes |
| Symptoms |  |  |  |  |
| (S1) Fever, with temperatures above 37.5 ° C for at least 3 consecutive days | -0.362 | 0.8421 | 0 | 0.80 |
| (S2) Cough | -0.426 | 0.810 | 0 | 0.81 |
| (S3) Sore throat and/or cold | -0.358 | 0.622 | 0 | 0.64 |
| (S4) Headache | -0.470 | 0.780 | 0 | 0.83 |
| (S5) Pain in muscles, bones, joints | -0.505 | 0.959 | 0 | 0.97 |
| (S6) Loss of taste and/or smell | -0.326 | 1.133 | 0 | 0.97 |
| (S7) Respiratory difficulty (sense of breathlessness at rest) | -0.246 | 1.305 | 0 | 1.03 |
| (S8) Chest pain (sternum pain) | -0.232 | 1.388 | 0 | 1.07 |
| (S9) Tachycardia (palpitations) | -0.209 | 1.374 | 0 | 1.05 |
| (S10) Gastrointestinal complaints (diarrhoea, nausea, vomiting) | -0.393 | 1.042 | 0 | 0.95 |
| (S11) Conjunctivitis (red eyes) | -0.164 | 1.170 | 0 | 0.88 |

There was no significant difference in the EPICOVID19 DS mean score between men (2.34±2.2) and women (2.49±2.4), while a low negative correlation between the score and the age of the participants was found (Spearman rho: −0.126; <0.001). The screening properties of the EPICOVID-19 DS compared to COVID-19 positive molecular and serological diagnosis are shown in table 4. The best value of Youden Index was observed for EPICOVID19 DS with respect to COVID-19 diagnosed by NPS(+). A good trade-off between sensitivity and specificity was observed (Se 76.56; Sp 68.24 AUC 77.5; 95% CI: 75.6-79.4). The cut-off score obtained was 2.56. Sensitivity and specificity improved when EPICOVID-19 DS was compared to COVID-19 detected by Antibodies IgG(+) (Se 80.37; Sp 80.17 AUC 86.0; 95% CI: 82.3-89.5) and the cut-off obtained was the same as for NPS(+) (2.59). The positive and negative predictive value for the IgG serological test was better than that for the nasopharyngeal swab (PPV IgG 54.43%: NPV IgG 93.27%). Finally, we observed a poor performance of the IgM results and are not presented in the table.

**Table 4.** Sensitivity and specificity of EPICOVID19 DC compared to COVID-19 positive molecular and serological diagnosis.

| Tested for SARS-CoV-2 |  |  |  |  |
| --- | --- | --- | --- | --- |
| Statistic | NASOPHARYNGEAL SWAB (2703)<br>(NPS+ 694 ) |  | ANTIBODIES IgG<br>(472)<br>(IgG+ 108 ) |  |
|  | Value | 95% CI | Value | 95% CI |
| Sensitivity | 76.56% | 72.99% to 79.87% | 80.37% | 71.58% to 87.42% |
| Specificity | 68.24% | 66.16% to 70.28% | 80.17% | 75.69% to 84.14% |
| Positive Likelihood Ratio | 2.41 | 2.23 to 2.61 | 4.05 | 3.23 to 5.08 |
| Negative Likelihood Ratio | 0.34 | 0.30 to 0.40 | 0.24 | 0.17 to 0.36 |
| COVID-19 Disease % | 23.29% | 21.68% to 24.96% | 22.77% | 19.05% to 26.83% |
| Positive Predictive Value | 42.26% | 40.38% to 44.17% | 54.43% | 48.77% to 59.98% |
| Negative Predictive Value | 90.55% | 89.23% to 91.74% | 93.27% | 90.40% to 95.33% |
| Accuracy | 70.18% | 68.39% to 71.93% | 80.21% | 76.32% to 83.72% |

## Discussion

Our focus was on developing a tool composed of simple questions related to the COVID-19 symptomatology for the identification of the subjects who are more likely to have SARS-CoV-2 infection in the general population. We validated the EPICOVID19 DS in a sample of voluntary subjects with serological and molecular clinical diagnosis. The optimal score, computed in 2703 adults aged 18 to 84 years, discriminates among symptomatic individuals. Before constructing the score, we performed both exploratory and confirmatory factor analysis to determine the number of factors/dimensions underlying the questionnaire. The results of these analyses supported a one factor model and the uni-dimensionality of the EPICOVID19 questionnaire. The magnitude of all factor loading was satisfactory, showing the highest factor loading value for the Respiratory difficulty, Chest pain, Tachycardia (palpitations) and Loss of taste and/or smell, Gastrointestinal complaints items appeared to be the most essential features of the EPICOVID19 DS. The high value for Chest pain is also explained by the fact that several patients reported it, possibly because of a tracheal pain caused by pneumonia^11^. Several clinical studies on hospitalised patients have shown that, at the onset of COVID-19, patients frequently show typical symptoms of viral pneumonia^12^. Symptoms that are less common, but still reported by a substantial number of patients are nasal congestion, sore throat, Gastrointestinal complaints and olfactory and taste disorders (OT-D)^13–15^. Subjects often reported Gastrointestinal complaints, not as isolated symptomatology of SARS-CoV-2 infection but as concurrent symptoms^16^. The lowest factor loading value was observed for the Sore throat and/or cold and Conjunctivitis. These lower values may be related to the fact that Conjunctivitis and cold were not the most frequent symptoms of COVID-19^17^. In line with other recent works^18,19^, the features encountered showed various aspects of the COVID-19 diagnosis definition. Indeed, Cough, loss of Taste and/or Smell and Respiratory difficulty were among the most reported symptoms in previous researches and corresponded to the items that gained the most importance in our score ^11,15,20,21^. However, the clinical presentation of COVID-19 disease is varied, and discrepancy may exist between symptoms and disease. A recent meta-analysis of symptoms including 50000 COVID-19 patients, found that fever and cough were the most common symptoms^22^ (89.1% and 72.2%) and a separate study of hospitalized subjects has suggested that respiratory distress has been reported in the most critical cases of COVID-19^23^. With the aim of supporting medical decision making, predicted models were developed for detecting people in the general population at risk of being admitted to hospital and for diagnosis of COVID-19 in patients with symptoms, but the results presented in a recent systematic review describe a poor research performance and a high risk of bias^24^. Using Homogeneity Analysis, we propose a scoring methodology for developing an improved scale. Therefore, we achieved a numerical weight value (optimal quantification) which represents the importance of the binary response categories (YES/NO) for each question of EPICOVID19 DS. As a result, the various binary items of the eleven questions of EPICOVID19 DS contributed with different weights to the overall score. This produced an improved scale, 0 to 10, reflecting the importance for each symptom. Thus, Respiratory problems and Chest pain were the most important symptoms, with score 1.03 and 1.07 respectively. The other symptoms showing an important contribution to the total score were Gastrointestinal complaint (0.95), Loss of Taste and/or Smell (0.97), Tachycardia (palpitations) (1.05). Subsequently, we computed sensitivity (Se) and specificity (Sp) of EPICOVID19 DS compared to COVID-19 positive serological and molecular diagnosis subjects. In NPS(+) subjects the cut-off score was 2.56 with sensitivity equal to 76.56% and specificity equal to 68.24%. In IgG(+) subjects the cut-off score was 2.59 and we obtained a substantial improvement of sensitivity, specificity, PPV and NPV with respect to NPS(+) subjects (Se 80.37; Sp 80.17; PPV 54.43%; NPV 93.27%). The COVID-19 sensitivity and specificity of serological and molecular diagnostic tests is not fully resolved but some studies suggest the sensitivity could be as low as 80%^25,26^. This raises concerns for a high false negative rate, which could result in an increase in the infection spread in the community. There is no absolute answer on the sensitivity and specificity of COVID-19 diagnostic tests because, to determine their accuracy, they must be compared with a “gold standard” test that currently is not identified. Considering estimates of Se and Sp, Positive Predictive Value (PPV) and Negative Predictive Value (NPV) can be gained on the basis of the disease prevalence and the rate of illness in the population but, as known, concerning COVID-19 there is significant uncertainty about this prevalence^27^. Statistically it is assumed that PPV varies widely, in a range between 30-50% in areas with low prevalence, as stated in a recent US research about COVID-19^28^. Early recognition screening and rapid diagnosis are essential to prevent transmission and provide supportive care in a timely manner. Nevertheless, screening is distinguished from further, more detailed diagnostic test assessment. This is of particular relevance as resources for full testing remains as a limited resource and optimizing its use is critical. EPICOVID19 DS could be a preliminary assessment that attempts to detect subjects with symptoms potentially associated with COVID-19 among a wide population. EPICOVID19 DS does not enable a clinical interview to determine the complete symptomatic profile and needs but identifies those who may warrant further assessment. An advantage would be to use this screening in primary care settings, so that GPs can avoid people with suspected COVID-19 in primary care’s offices when possible^29^, or as a first screening tool and then manage the patient remotely by telephone or video consultations^30^. The EPICOVID19 DS could be applied to the general population. Once the scoring is assigned to each symptom, the EPICOVID19 DS could allow to set different cut-offs according to the subjects involved and to the gold standards used (NPS, serological tests, clinical evaluation by clinicians, etc.). It should be noted that since it is plausible to expect lower prevalence values in the general population than the 22.77% of the present series, the probability of NPV would increase beyond the current 93.27% and consequently the probability of progressing to COVID-19 for subjects who tested negative (1 - NPV) would be less than the current 6.7%. Although the identified symptoms are not specific for COVID-19, they have been found as valid references in a population setting because they are frequently reported by patients affected by COVID-19. Likely, in a non-pandemic scenario, these symptoms could be assessed with different weights because of their aspecificity, configuring the EPICOVID19 DS as a valid diagnostic support mainly in a pandemic situation. Moreover, the health authority is still unable to monitor through classic tests the spread of SARS-CoV-2 infection, and allowing the circulation of unsuspecting positive subjects could represent a risk for the spread of the infection. The validation of an instrument that can easily identify a suspected case, through a score attributed to each symptom related to COVID-19, can be of great importance in facilitating the containment of the epidemic. The proposed score seems worthy of validation in broader populations in order to confirm its clinimetric properties. In the event of a confirmatory answer, it might qualify as a useful means of selecting people amenable to serological and/or molecular diagnostic tests for COVID-19. The availability and offering of diagnostic tests for the SARS-CoV-2 coronavirus has proven to be one of the keys to the containment of the COVID-19 pandemic. The early identification of positive subjects at molecular and serological tests among people with specific symptoms or considered at risk is crucial to limit the spread of the infection. The tool we validated responds to the need to readily identify a suspect case, through a score attributed to each symptom related to COVID-19. Although the validation was satisfactory, the proposed score seems worthy of being further testing in larger populations in order to confirm its clinimetric properties, useful for selecting people susceptible to serological and / or molecular diagnostic tests for COVID-19.

Although this tool could be a public health prevention measure instrument, directing subjects to a self-assessment without trigger panic, alarmism and concern among the screened population, some limitations need to be outlined. First of all, the participation in the study was voluntary and not representative of the general population. This presents potential selection biases that must be taken into consideration. Data were collected in a convenient young-adult and highly educated population sample characterized by low multimorbidity, as resulting from the phase I study^9^ and expectable in the case of an on-line questionnaire promoted by e-mail invitations. Further, in the context of a pandemic, our survey might have interested people who had no opportunity to report symptoms to clinicians. Moreover, the effect linked to recall bias cannot be excluded among the participants who tested positive at COVID-19 and/or presenting symptoms related to the SARS-COV2 infection. Given these limitations, the adoption of EPICOVID19 DS should be considered with caution and the procedures outlined for its development could be applied iteratively as new data is collected to continue refinement of this potentially valuable clinical decision support tool.

## Data Availability

All manuscript data are available on request

## Contributors

SM, LB, FA and FP were responsible for the study concept and design. LB, LF and SP were responsible for the literature search. FA, LF and SP were responsible for acquisition of data. LB, FA, FB, FP and SM were responsible for analysis and interpretation of data. LB, LF, SM, SP and FB were responsible for drafting of the manuscript. MG, AG, RAI, CP, MA and GP were responsible for critical revision of the manuscript for important intellectual content. LB, SM were responsible for statistical analysis. GP, SM, CT, MN, NJ, AS, CP, MA critically revised the manuscript for important intellectual content. All authors participated in data interpretation, read, and approved the final version. The corresponding author, SM, attests that all listed authors meet authorship criteria and that no others meeting the criteria have been omitted.

## Ethical statement and consent form

EPICOVID19 phase II study was approved by the Ethical Committee of the Istituto Nazionale per le Malattie Infettive I.R.C.C.S. ‘L. Spallanzani’ as an amendment of the EPICOVID19 epidemiological study (approval No. 93 of the trial register). Data transfer was safeguarded by means of password protection and encrypting/decrypting policy; all data were handled and stored in accordance with the EU GDPR 2016/679 (http://gdpr-info.eu/). Informed consent was accessible on the home page of the platform and participants were asked to review before starting the compilation, thus explaining the purpose of the study and which data were to be collected and how data were stored.

Email address is the personal data provided on a voluntary basis in Phase I. In our study it was only used: 1) to send email invitation to the Phase II survey; 2) to link the information related to the results (swabs and/or antibodies IgG, IgM) to the information on symptoms collected during the phase I survey. In the participation mail, the person was able to participate by clicking on the provided link to the survey; to not participate ignoring the invitation; to communicate to a specific address the request of deletion of the email address from the database.

## Acknowledgments

We thank Prof. Mario Grassi of the University of Pavia for his suggestions in statistical analysis and Dr. Michael N. Liebman for his support in the final revision of the manuscript contents and their formalization in English.

To the Editor of Journal of Medical Internet Research (JMIR; IF 5.03)

Gunther Eysenbach, MD, MPH

Senior Scientist, Centre for Global eHealth Innovation

Tecnha Institute and Toronto General Research Institute of the UHN;

Dear Editor,

We would like to submit for publication our manuscript entitled: “EPICOVID19: Psychometric assessment and validation of a short diagnostic scale for a rapid Covid-19 screening based on reported symptoms “, authors: Luca Bastiani, Loredana Fortunato, Stefania Pieroni, Fabrizio Bianchi, Fulvio Adorni, Federica Prinelli, Andrea Giacomelli, Gabriele Pagani, Stefania Maggi, Caterina Trevisan, Marianna Noale, Nithiya Jesuthasan, Aleksandra Sojic, Carla Pettenati, Massimo Andreoni, Raffaele Antonelli Incalzi, Massimo Galli, Sabrina Molinaro.

This study represents a collaboration between Italian National Research Council (Institute of Clinical Physiology, Institute of Biomedical Technologies, Institute of Neuroscience) and three important Italian Universities: of Rome (Unit of Geriatrics), Padova (Department of Medicine) and Milano (Infectious Diseases Unit, Department of Biomedical and Clinical Sciences).

We opted-in to medRxiv preprint Review service for our paper on 22-07-2020 (ID manuscript MEDRXIV/2020/159590).

The aim of this work is to assess the capability of specific self-reported symptoms collected through the EPICOVID19 questionnaire to discriminate the COVID-19 among symptomatic subjects in order to identify possible COVID-19 cases, to undergo instrumental measurements and clinical examinations. Early recognition and rapid diagnosis are essential to prevent transmission and provide supportive care in a timely manner, crucial elements in a pandemic scenario to support public health and prevention policies.

## Evidence before this study

As of July 28 over 16 million cases of COVID-19 have been reported to WHO.

SARS-CoV-2 strain responsible for the epidemic in Italy entered the country at the end of January. Italy has been the one of the world’s worst affected countries both in terms of infections and deaths. Rapid screening tests and technological support are needed for outbreak control and surveillance. In Italy due to organizational difficulties not yet overcome, the test for determining the presence of viral RNA by RT-PCR on nasopharyngeal swab (NPS) was performed almost exclusively to people with moderate-to-severe respiratory symptoms. Despite the numerous requests made by the population for a dedicated emergency number, the restriction to the access to tests remained throughout the entire most active epidemic phase. It is therefore likely that thousands of people with mild to severe COVID-19 remained at home during the lockdown without being able to access a diagnostic test. In such a situation, the identification of symptoms that, considered together, allow to formulate a diagnosis of probable COVID-19, acquires particular relevance. Large-scale programs using testing and screening are currently under evaluation by different governments, but clear understanding of the optimal population and role for these tests in the care pathway, are currently lacking.

## Added value of this study

Our study proposes a short diagnostic scale (EPICOVID19 DS) to early recognize conditions attributable to the infection and for the timely identification of cases with possible unfavorable evolution. The use of the scale could be important in the epidemiological context for a rapid identification of suspected cases (with SARS-CoV-2 infection-like symptoms and signs) to be immediately subjected to quarantine measures. Our study provides a critical method for evaluating and interpreting self-reported data as part of COVID-19 patient management by development of a multi-component score.

## Implications of all the available evidence

Early recognition screening and rapid diagnosis are essential to prevent transmission and provide supportive care in a timely manner and our score supports the potential for identifying individuals who need to seek immediate clinical evaluation. In a pandemic scenario, EPICOVID19 DS could be a preliminary assessment that attempts to detect subjects with symptoms potentially associated with COVID-19 among a wide population.

All authors have read and approved the final version of the manuscript.

The paper has not been published before and is not under consideration for publication in whole or in part elsewhere. If accepted by Journal of Medical Internet Research (JMIR; IF 5.03), it will not be published elsewhere in the same form, either in English or in any other language, without the consent of the Editors.

Hoping to hearing from you soon,

Sincerely yours,

Sabrina Molinaro

Institute of Clinical Physiology, Italian National Research Council

Via Moruzzi 1, 56124, Pisa - Italy

## Reference

1 WHO. Coronavirus (COVID-19) events as they happen. 2020.https://www.who.int/emergencies/diseases/novel-coronavirus-2019 https://www.who.int/emergencies/diseases/novel-coronavirus-2019 (Accessed May 23, 2020).

2 Johns Hopkins University and Medicine. COVID-19 map. Johns Hopkins Coronavirus Resource Centre. https://coronavirus.jhu.edu/map.html (accessed April 12, 2020)

3 COVID-19 Clinical Research Coalition. Electronic address. Global coalition to accelerate COVID-19 clinical research in resource-limited settings. Lancet. 2020;395(10233):1322–1325. doi:10.1016/S0140-6736(20)30798-4

4 Lipsitch M, Swerdlow DL, Finelli L. Defining the epidemiology of Covid-19 - studies needed. N Engl J Med. 2020; 382: 1194–96.

5 WHO. Population-based age-stratified seroepidemiological investigation protocol for COVID-19 virus infection. March 17, 2020. https://apps.who.int/iris/handle/10665/331656 (accessed June 20, 2020).

6 Green et al. Molecular and antibody point-of-care tests to support the screening, diagnosis and monitoring of COVID-19, April 2020, Oxford University.

7 WHO. Global surveillance for COVID-19 caused by human infection with COVID-19 virus. Interim guidance 20 March 2020.

8 Reeves et al. Biblio Rapid response to COVID-19: health informatics support for outbreak management in an academic health system. J Am Med Inform Assoc. 2020 Jun 1; 27(6):853–859.

9 Adorni F, Prinelli F, et al. Self-reported symptoms of SARS-CoV-2 infection in a non-hospitalized population: data from the large Italian web-based EPICOVID19 cross-sectional survey. JMIR Preprints. 27/06/2020:21866 DOI: 0.2196/preprints.21866 URL: https://preprints.jmir.org/preprint/21866

10 Long Q, Liu B, Deng H, et al. Antibody responses to SARS-CoV-2 in patients with COVID-19. Nat Med. 2020 Jun; 26(6):845–848.

11 Chan JFW, Yuan S, Kok KH, et al. A familial cluster of pneumonia associated with the 2019 novel coronavirus indicating person-to-person transmission: a study of a family cluster. Lancet 2020; 395: 514–23.

12 Giacomelli A, Ridolfo AL, Milazzo L, et al. 30-day mortality in patients hospitalized with COVID-19 during the first wave of the Italian epidemic: A prospective cohort study. Pharmacol Res. 2020; 158:104931.

13 Giacomelli A, Pezzati L, Conti F, et al. Self-reported olfactory and taste disorders in SARS-CoV-2 patients: a cross-sectional study. Clin Infect Dis. 2020;ciaa330.

14 Pallanti Importance of SARs-Cov-2 anosmia: From phenomenology to neurobiology. May 2020; Compr Psychiatry. 2020 Jul; 100:152184.

15 Menni et al. Quantifying additional COVID-19 symptoms will save lives. Lancet 2020 Jun 20; 395(10241):e107–e108.

16 Cholankeril et al. High Prevalence of Concurrent Gastrointestinal Manifestations in Patients with SARS-CoV-2: Early Experience from California. Gastroenterology 2020 Apr 10; S0016-5085(20)30471-6.

17 Sarma P, Kaur H, Kaur H, et al. Ocular Manifestations and Tear or Conjunctival Swab PCR Positivity for 2019-nCoV in Patients with COVID-19: A Systematic Review and Meta-Analysis (3/30/2020). SSRN Electronic Journal. 2020 Apr 15. doi:10.2139/ssrn.3566161

18 Huang C, Wang Y, Li X, et al. Clinical features of patients infected with 2019 novel coronavirus in Wuhan, China. Lancet 2020 Feb 15; 395(10223):497–506.

19 Guan WJ, Ni ZY, Hu Y, et al. Clinical characteristics of coronavirus disease 2019 in China. N Engl J Med. 2020 Apr 30; 382(18):1708–20.

20 Huang C, Wang Y, Li X, et al. Clinical features of patients infected with 2019 novel coronavirus in Wuhan, China. Lancet 2020; 395: 497–506.

21 Bénézit F, Le Turnier P, Declerck C, et al. Utility of hyposmia and hypogeusia for the diagnosis of COVID-19. Lancet Infect Dis. 2020 Apr 15. pii: S1473-3099(20)30297-8.

22 Sun P, Qie S, Liu Z, Ren J, Li K, Xi J. Clinical characteristics of hospitalized patients with SARS-CoV-2 infection: A single arm meta-analysis. J Med Virol. 2020 Feb 28. 92(6):612–617.

23 Jain V, Yuan J-M. Systematic review and meta-analysis of predictive symptoms and comorbidities for severe COVID-19 infection. medRxiv 2020; published online March 16. DOI: https://doi.org/10.1101/2020.03.15.20035360

24 Wynants L, Van Calster B, Bonten MMJ, et al. Prediction models for diagnosis and prognosis of covid-19 infection: systematic review and critical appraisal. BMJ 2020 Apr 7; 369:m1328

25 Wang W, Xu Y, Gao R, et al. Detection of SARS-CoV-2 in different types of clinical specimens. JAMA 2020; 323(18):1843–1844

26 Yong SEF, Anderson DE, Wei WE, et al. Connecting clusters of COVID-19: an epidemiological and serological investigation. Lancet Infect Dis. 2020 Apr 21. 20(7):809–815.

27 Charles F. Manski Bounding the Predictive Values of COVID-19 Antibody Tests. medRxiv 2020; published online May 18. DOI: https://doi.org/10.1101/2020.05.14.20102061

28 Mathur G, Mathur S. Antibody testing for COVID-19: Can it be used as a screening tool in areas with low prevalence? Am J Clin Pathol 2020.;154:1–3.

29 Razai MS, Doerholt K, Ladhani S, et al. Coronavirus disease 2019 (covid-19): a guide for UK GPs. BMJ 2020 Mar 5; 368:m800.

30 Greenhalgh T, Koh GCH, Car J. Covid-19: a remote assessment in primary care. BMJ 2020 Mar 25; 368:m1182.

